# SARS-CoV-2 RNA wastewater settled solids surveillance frequency and impact on predicted COVID-19 incidence using a distributed lag model

**DOI:** 10.1101/2022.02.21.22270864

**Authors:** Mary E. Schoen, Marlene K. Wolfe, Linlin Li, Dorothea Duong, Bradley J. White, Bridgette Hughes, Alexandria B. Boehm

## Abstract

SARS-CoV-2 RNA concentrations in wastewater settled solids correlate well with COVID-19 incidence rates (IRs). Here, we develop distributed lag models (DLMs) to estimate IRs using concentrations of SARS-CoV-2 RNA from wastewater solids and investigate the impact of sampling frequency on model performance. SARS-CoV-2 N gene and PMMoV RNA concentrations were measured daily at four wastewater treatment plants in California. Artificially reduced datasets were produced for each plant with sampling frequencies of once every 2, 3, 4, and 7 days. Sewershed-specific models that related daily N/PMMoV to IR were fit for each sampling frequency with data from mid-Nov 2020 through mid-July 2021, which included the period of time during which Delta emerged. Models were used to predict IRs during a subsequent out-of-sample time period. When sampling occurred at least once every 4 days, the in- and out-of-sample root mean square error (RMSE) changed less than 7 cases/100,000 compared to daily sampling across sewersheds. This work illustrates that real-time, daily predictions of IR are possible with small error, despite changes in circulating variants, when sampling frequency is once every 4 days or more. However, reduced sampling frequency may not serve other important wastewater surveillance use cases.

## 1.0 Introduction

Wastewater represents a pooled biological sample from the contributing community and is a resource for assessing population health. Wastewater-based epidemiology has been used to assess infectious disease occurrence^1–3^ and identify high rates of substance abuse.^4,5^ The COVID-19 pandemic has greatly increased interest in utilizing wastewater-based epidemiology to supplement clinical testing data, which can be limited due to test seeking behavior and test availability.^6^ Throughout the pandemic, researchers have successfully detected and monitored SARS-CoV-2 RNA in wastewater^7–11^ and programs have been developed to aid public health decision makers in assessing the occurrence of COVID-19 in their communities.^12,13^ SARS-CoV-2 RNA concentrations in wastewater solids have been used in various modeling approaches to assess or predict COVID-19 case counts or incidence rates. Mechanistic modeling approaches estimated cases from wastewater settled solids using a mass balance approach that included as inputs the SARS-CoV-2 RNA from shedders and its fate and transport in the sewer system.^14^ Alternatively, regression approaches utilizing daily wastewater data indicated strong stochastic relationships between gene concentration and cases or incidence without additional inputs such as shedding rate.^8,15^ The motivation of this work was to determine if wastewater surveillance could retain the benefit of real-time prediction of incidence rates using a regression approach when scaled to sustainable and cost-effective sampling frequencies.

Distributed lag models (DLMs), in which the effect of an explanatory variable on the dependent variable occurs over time rather than all at once, have been applied previously to investigate the relationship between SARS-CoV-2 RNA concentrations in wastewater solids and laboratory confirmed COVID-19 incident cases.^10,16^ DLMs are most useful for prediction when explanatory and dependent variables are stationary (i.e., the probability distribution is time independent).^17^ For SARS-CoV-2 RNA wastewater surveillance, there is an interest in models that perform well for prediction both when sampling is conducted at intervals that are less burdensome than daily and as the virus acquires mutations that may affect characteristics of infection that alter observations in wastewater such as shedding and transmissibility.

We utilize daily SARS-CoV-2 RNA concentrations in wastewater settled solids to evaluate a set of DLMs using out-of-sample prediction and artificially reduced sampling frequency during a time of substantial changes in levels of community transmission and circulating variants across four sewersheds. We assess the impact of sampling frequency on prediction error to support robust methods of COVID-19 incidence rate prediction using reduced sampling across a range of sewersheds with varying sizes and characteristics. Inherent in this analysis is the assumption that the desired use case for wastewater SARS-CoV-2 surveillance is development and implementation of a real-time model for predicting laboratory-confirmed incidence rates across a wide range of changes in infection dynamics. Although DLM have been applied before to model COVID-19 cases using wastewater,^10,16^ they have not been used to identify ideal sampling frequencies nor have they been used to investigate whether emergence of a new variant alters the relationship between cases and wastewater concentrations of SARS-CoV-2 RNA.

## 2.0 Methods

The Institutional Review Board of Stanford University determined that this project does not meet the definition of human subject research as defined in federal regulations 45 CFR 46.102 or 21 CFR 50.3 and indicated that no formal IRB review is required.

### 2.1 Wastewater data

Wastewater settled solids samples were collected daily at four POTW as part of a regional monitoring program for SARS-CoV-2 RNA in wastewater. Two POTWs serve populations of Santa Clara County, CA (SJ and PA), and the others each serve a portion of San Mateo County (PA), Sacramento County (Sac), and Yolo County (Dav). The four POTWs serve between 66,000 and 1,500,000 residents and have permitted flows between 7.5 and 181 million gallons per day. Further details of the POTWs are provided in Wolfe et al.^15^

Samples were collected by POTW staff using sterile technique in clean, labeled bottles. POTWs were not provided compensation for participation. Approximately 50 ml of settled solids was collected each day from each POTW between mid-to-late November 2020 and September 2021.

Settled solids were collected from the primary clarifier; at these POTWs, the residence time of solids in the primary clarifier ranged from 1 to 3 h.^15^ Settled solids samples were grab samples at all plants except for SJ. At SJ, POTW staff manually collected a 24-h composite sample.^8^ Samples were immediately stored at 4°C and transported to a commercial partner laboratory by a courier service where processing began within 6 h of collection.

At the laboratory, the concentration of the SARS-CoV-2 N gene and PMMoV RNA were measured, along with recovery of an external control (bovine coronavirus, BCoV) using a high-throughput method that used liquid handling robots and automated equipment. The methods used to quantify these RNA targets are explained in detail in Wolfe et al.^15^ and protocol.io,^18^ so they are not repeated herein. In brief, solids were dewatered using a centrifuge and then suspended in a buffer and homogenized. RNA was extracted and purified from the buffer and inhibitors were removed using commercial kits. RNA targets were quantified using digital droplet RT-PCR, and concentrations of the N gene and PMMoV are expressed as copies per g dry weight of solids. The ratio of N/PMMoV was then calculated for use in this study. BCoV recovery had to be higher than 10% for the sample, or the sample was rerun. Positive and negative controls had to be positive and negative, respectively, or the sample was rerun. All samples were run in 10 replicates. Data collected from the start of the project through 31 March 2021 are published in a peer-reviewed paper by Wolfe et al.^15^ The data from 1 April 2021 through September 2021 have not previously been published, and the analysis presented herein was not previously completed on any of the included data. Wastewater data are available publicly in the Stanford Digital Repository (https://purl.stanford.edu/gg221yy5416).

The above analysis resulted in N/PMMoV once per day for each of the four POTW. We downsampled the daily data to obtain data at 4 additional lower sampling frequencies at each POTW: once every 2 days, once every 3 days, once every 4 days, and weekly. The sub-daily frequency datasets were generated from the daily N/PMMoV datasets for model fitting by artificially removing observations starting with the second observation.

### 2.2 COVID incidence data

Laboratory-confirmed COVID-19 incident cases as a function of episode date (earliest of reported symptom onset, laboratory result, or case record create dates) for each sewershed were obtained from local or state sources. Case data were aggregated within the sewersheds based on georeferenced reported home addresses, which were delineated using the POTW-specific geographic information system (GIS) shape files. COVID-19 incidence rates were calculated using the estimated population served in each sewershed. 7-day centered smoothed incidence rates (hereafter, IRs) were used as the dependent variable. The smoothing reduced the “weekend effect” associated with a reduction in test seeking behavior, testing availability, and result reporting.^19^

### 2.3 Estimated percent of cases caused by the Delta variant

The percent of total monthly cases resulting from the Delta variant for the state of California were obtained from publicly available data from the CA Department of Public Health.^20^ The state data was used as an explanatory variable in regression models for each of the four POTW in the absence of site-specific data. Linear interpolation was used to estimate the daily percentage of cases associated with Delta (% Delta) with the monthly value assigned to the beginning of each month.

### 2.4 Model

#### 2.4.1 Model candidates: daily data

We fit a variety of regression models using the daily sampling dataset for each POTW: a linear model (Eq 1); DLMs (Eq 2); and variations of Eq 2 with % Delta added as an explanatory variable (Eqs 3-4):

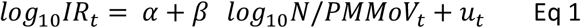

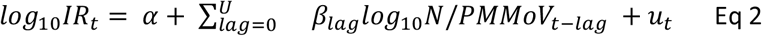

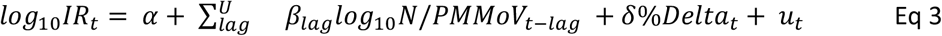

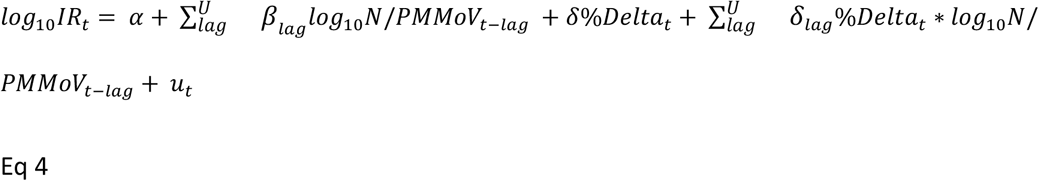

where *IR*_*t*_ is the smoothed 7-day incidence rate at time t (unitless); *lag* is the number of days before the prediction date (time t); *U* is the upper limit of lag; *N/PMMoV*_*t*-*lag*_ is the ratio of N/PMMoV at time t-lag; *α* is a constant indicating the intercept; %*Delta*_*t*_ is the percentage of laboratory-confirmed COVID-19 cases associated with the Delta variant at time t; *β, β*_*lag*_, *δ*, and *δ*_*lag*_ are coefficients; and *u*_*t*_ are error terms, which are assumed to be independently and identically distributed. Eq 2 was fit assuming an upper lag limit *U* of 3 to 6; *U* of 3 was selected to match with the earliest day used to calculate the smoothed IR (see Section 2.2). *U* of 6 was arbitrarily selected and the impact of additional predictors was explored using Bayesian Information Criterion (described in Section 2.4.3). Eqs 3-4 explore possible impacts of the Delta variant on incidence rate without and with an interaction term, respectively, using the lags from Eq 2. The results of this analysis informed the models investigating for use with a reduced wastewater data frequency.

#### 2.4.2 Model candidates: reduced sampling frequency

We fit both a linear model (Eq 1) and DLM (Eq 5) to each reduced sampling frequency dataset for each POTW. A unique DLM with two predictors was fit for each sampling frequency:

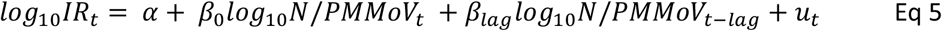

See Eq 2 for regressor descriptions. The lag was determined by the frequency of sampling (i.e. every other day had a lag of 2; once every 3 days had a lag of 3; once every 4 days had a lag of 4 and weekly had a lag of 7).

#### 2.4.3 Model fit computation and selection criteria

IR was modeled as a function of N/PMMoV for each sampling frequency and POTW using the in-sample time period of 11/15/20 – 7/19/2021, with one exception. Models in Eq 3-4 were fit using the entire dataset to capture the full range of % Delta. The model parameters were estimated by ordinary least square (OLS) method using the Dynlm function in R.^21^ SARS-CoV-2 N gene non-detects were modeled as half the limit of detection of 500 gene copies/g dry weight.

One model was selected from the candidates for each sampling frequency. Model selection considered first the interpretation and statistical significance of coefficient estimates across POTWs. The resulting candidate models for each sampling frequency were compared by Bayesian Information Criterion (BIC). In a model with K coefficients, including the intercept, the BIC was calculated:

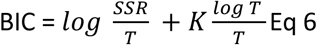

where SSR is the sum of the squared residuals and T is the number of observations. A difference in BIC>1 indicated a preference for the model with the smallest value when predicting the same set of IR observations. In addition, the in-sample residuals were tested for auto-correlation using ACF function in R.

#### 2.4.3 Model performance

For the selected models, the root mean square error (RMSE) was calculated for the in-sample time period of 11/15/20 – 7/19/2021 and out-of-sample period of 7/20/21 – 9/15/2021, separately, and reported in terms of IR as (cases/100,000). For all sampling frequencies, the predicted IR on days with missing data (either artificially removed or in the daily sampling dataset) were set to the previous predicted IR to compare performance across sampling frequencies using the same set of observations. In addition to the reduced sampling frequency N/PMMoV datasets generated to fit the models, additional datasets were generated with staggered starting dates to capture the possible variation resulting from sampling at reduced frequency. This resulted in one N/PMMoV dataset for once per day sampling, 2 datasets for once every other day sampling and so forth. The minimum, median, and maximum RMSE was reported for each sampling frequency. The median out-of-sample RMSE was compared to the median in-sample RMSE for each POTW and sampling frequency for out-of-sample model validation.

#### 2.4.4 Comparison of sampling frequency for prediction

Whereas the in-sample and out-of-sample RMSE were compared for model validation, the out-of-sample RMSEs were compared across sampling frequency for each POTW to evaluate the effect of sampling frequency on accuracy of prediction. Because the highest IR peak occurred during the in-sample period, the in-sample RMSEs were also compared across sampling frequency to evaluate the models across the full range of observed IR.

## 3.0 Results

### 3.1 Measurement Overview

During the sampling period of 11/15/20 – 9/15/2021, the range of SARS-CoV-2 RNA N gene concentrations was non-detect to 3.71 × 10^6^ cp/g and the range of PMMoV concentrations was 7.12 × 10^7^ to 3.74 × 10^10^ cp/g across the four POTWs. There were a total of 0, 9, 0, and 10 non-detects at SJ, PA, SAC, and Dav, respectively. Data were missing on 3/23/2021 and 9/12/2021 at PA; 1/20/21 and 5/25/2021 at Sac; and 2/14/21, 2/15/21 and 7/20/21 at Dav. The resulting N/PMMoV range was non-detect to 1.25 × 10^−2^. The maximum and minimum N/PMMoV and (SARS-CoV-2 RNA N gene concentrations) occurred before the Delta variant surge (Figure 1).

**Figure 1.**
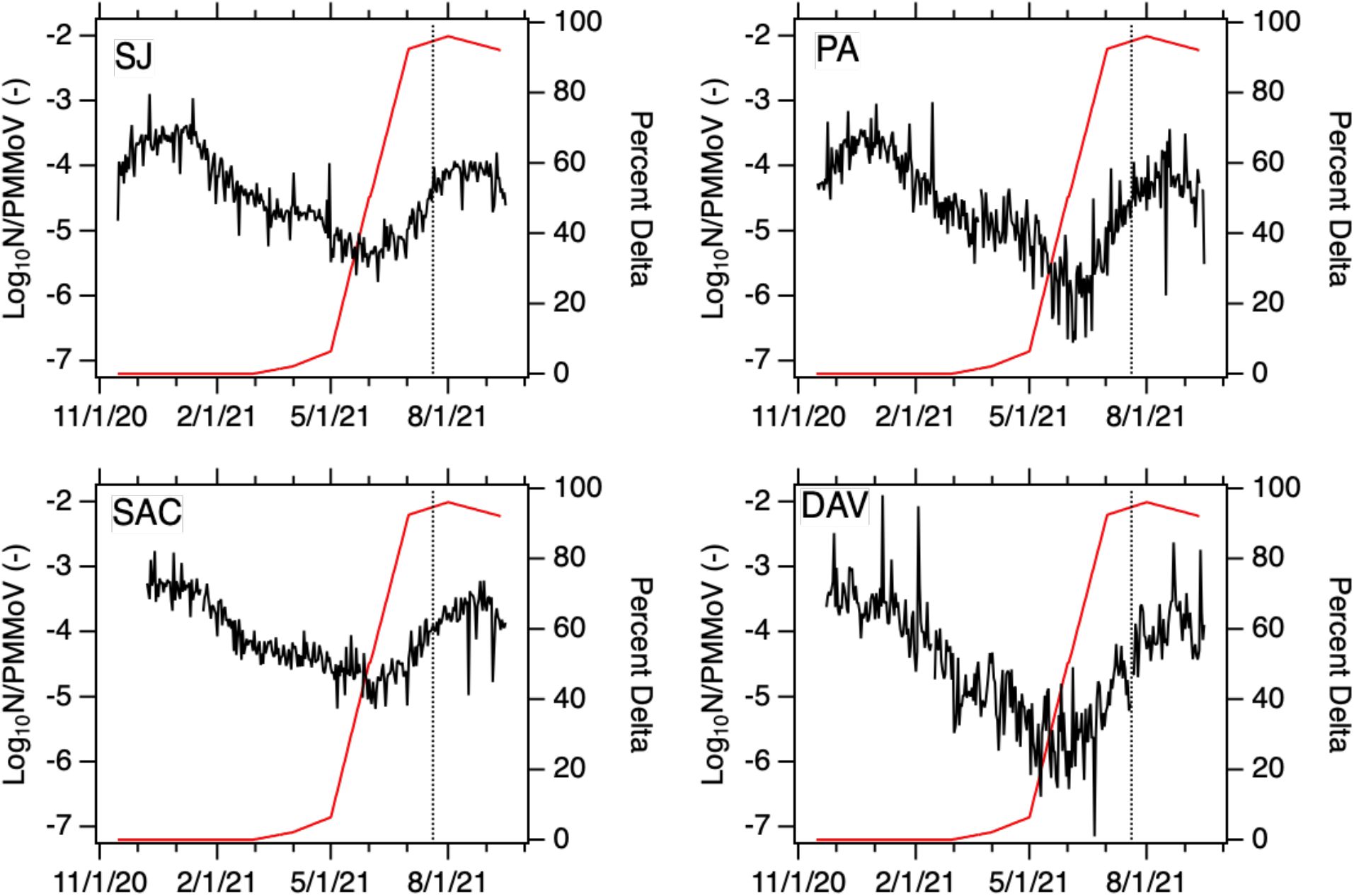
Log_10_ N/PMMoV by POTW (black) and the percentage of clinical samples that are the Delta variant (% Delta, red) in California. Vertical black line indicates beginning of out-of-sample time period (20 July 2021 until end of data).

### 3.2 Model selection

#### 3.2.1 Daily sampling frequency

We first selected the best performing model from the candidate models (Eqs 1-4) fit using the daily N/PMMoV sampling data available from the four POTWs during the in-sample period of 11/15/20 – 7/19/2021 for Eqs 1-2 and entire sampling period 11/15/20 – 9/15/2021 for Eqs 3-4. Tables S1-S4 present model fit results for the candidate models including the BICs. DLMs that incorporated % Delta as a predictor (Eqs 3-4) were eliminated due to coefficient estimates for % Delta and the interaction terms that were not statistically different from zero (p < 0.1) (not shown). The selected model (Eq 2 with U=3; model fit presented in Table 1 for SJ) was preferable to the linear model (Eq 1) based on the BIC across the four POTWs. The inclusion of additional predictors (with U of 4, 5, 6) was not supported by the BIC (Tables S1-4). The coefficient estimates of the explanatory variables for Eq 2 with maximum lag of U = 3 were significantly different from 0 across all POTWs (p < 0.01) and positive (Table 1 and Table S1-S4), indicating that an increase in log_10_ N/PMMoV was associated with an increase in log_10_ IR. The selected model was used to investigate out-of-sample prediction given daily wastewater settled solids data.

**Table 1.** SJ model fit for dependent variable log_10_ IR (i.e., log_10_ 7-day smoothed cases/population)

| Predictor | Coefficient Estimate for<br>predictors (St. Error) |  |  |  |  |
| --- | --- | --- | --- | --- | --- |
|  | Daily | Once<br>every 2<br>days | Once<br>every 3<br>days | Once<br>every<br>4 days | Weekly |
| $\log_{10}$<br>N/PMMoV<sub>t-0</sub> | 0.25***<br>(0.03) | 0.48***<br>(0.04) | 0.52***<br>(0.04) | 0.53***<br>(0.06) | 0.63***<br>(0.10) |
| $\log_{10}$<br>N/PMMoV<sub>t-1</sub> | 0.22***<br>(0.03) | | | | |
| $\log_{10}$<br>N/PMMoV<sub>t-2</sub> | 0.23***<br>(0.03) | 0.40***<br>(0.04) | | | |
| $\log_{10}$<br>N/PMMoV<sub>t-3</sub> | 0.22***<br>(0.03) | | 0.36***<br>(0.04) | | |
| $\log_{10}$<br>N/PMMoV<sub>t-4</sub> | | | | 0.35***<br>(0.06) | |
| $\log_{10}$<br>N/PMMoV<sub>t-7</sub> | | | | | 0.34***<br>(0.10) |
| $\alpha$ | 0.03<br>(0.05) | -0.13<br>(0.09) | -0.07<br>(0.08) | -0.07<br>(0.12) | 0.30<br>(0.24) |
| Adjusted R<sup>2</sup> | 0.96 | 0.94 | 0.97 | 0.95 | 0.91 |

\* p<0.1; \*\* p<0.05; \*\*\* p<0.01

#### 3.2.2 Reduced sampling frequency

We also fit linear and DLMs (Eq 1 and Eq 5) using the reduced sampling frequency wastewater data for the four POTWs. Table 1 presents example DLM in-sample model fit results for SJ (Tables S5-S7 present in-sample model fit results for Dav, PA, and Sac). The DLMs (Eq 5) were selected over the linear model for each sampling frequency based on the higher R^2^ values in-sample; the difference in BIC was < 1.0 between the DLMs and linear models, indicating no preference (results not shown). The coefficient estimates for the regressors were significantly different from 0 (p<0.01) (with the exception of the Sac weekly model) and positive.

### 3.3 Coefficient estimates

DLM coefficient estimates (Table 1) indicate the effect of a temporary change in log_10_ N/PMMoV at time t on log_10_ IR in period t + lag. For the daily sampling DLM, the immediate impact effect (t=0) was greatest for all POTWs. The coefficient estimates of the lags often fell within the standard error of the coefficient estimate of t=0 (Table 1 and Figure 2). For DLMs fit using reduced sampling frequency, the immediate impact effect (t=0) was greatest only for SJ (Table 1 and Tables S5-S7). For Dav, PA, and Sac, the immediate impact effect was less than the lag impact effect across reduced sampling frequencies. Across POTWs, whereas the linear regression coefficients for N/PMMoV for daily sampling had a wide range (0.51 - 0.84), the coefficient estimates for the DLMs for daily sampling were similar with overlapping standard errors; however, as sampling frequency decreased, the coefficient estimates of the DLMs for each POTW diverged (Figure 2).

**Figure 2.**
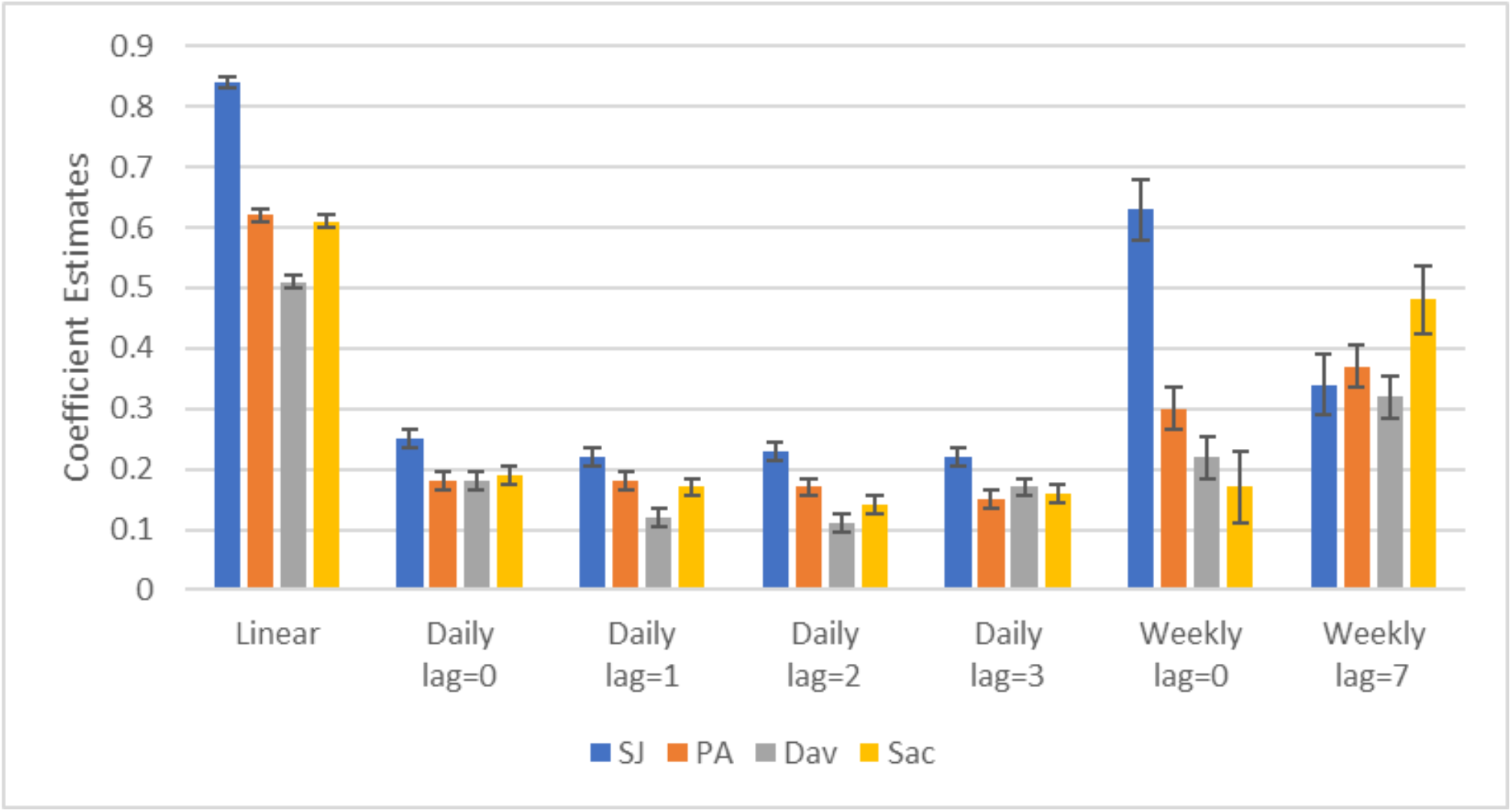
Coefficient estimates for the linear model and the DLMs for daily sampling and weekly sampling with corresponding standard errors (error bars).

### 3.4 Model performance

The out-of-sample model performance was evaluated by comparing the in- and out-of-sample RMSEs. For the models trained with daily data, the out-of-sample RMSE was less than the in-sample RMSE for SJ and PA with an increase of roughly 3 cases/100,000 for Dav, indicating good out-of-sample model performance (Figure 2). The Sac out-of-sample RMSE was greater than the in-sample RMSE by roughly 10 cases/100,000.

For models fit using wastewater data collected at reduced sampling frequencies (between once every 2 to 7 days), the RMSE was calculated by freezing the IR prediction at the previous predicted value for days when there was no data both in the in-sample and the out-of-sample time periods. The median out-of-sample RMSEs for models fit using the reduced sampling frequency wastewater data were at most 3 cases/100,000 greater than the median in-sample RMSE for each sampling frequency across POTWs, with the exception of Sac with a maximum difference of roughly 7 cases/100,000.

The predicted IR traces (Figure 3 for SJ and Figures S1 for PA, Dav, and SAC) show that the DLMs captured the in-sample and out-of-sample surges in reported IR across POTWs. The daily DLMs generally underpredicted IR during peak periods (residual plots presented in Figures S2-S5). There is considerable variation in prediction when predicting IR using reduced sampling frequency depending on the specific days that are sampled (weekly prediction is presented in Figure 4 for SJ).

**Figure 3.**
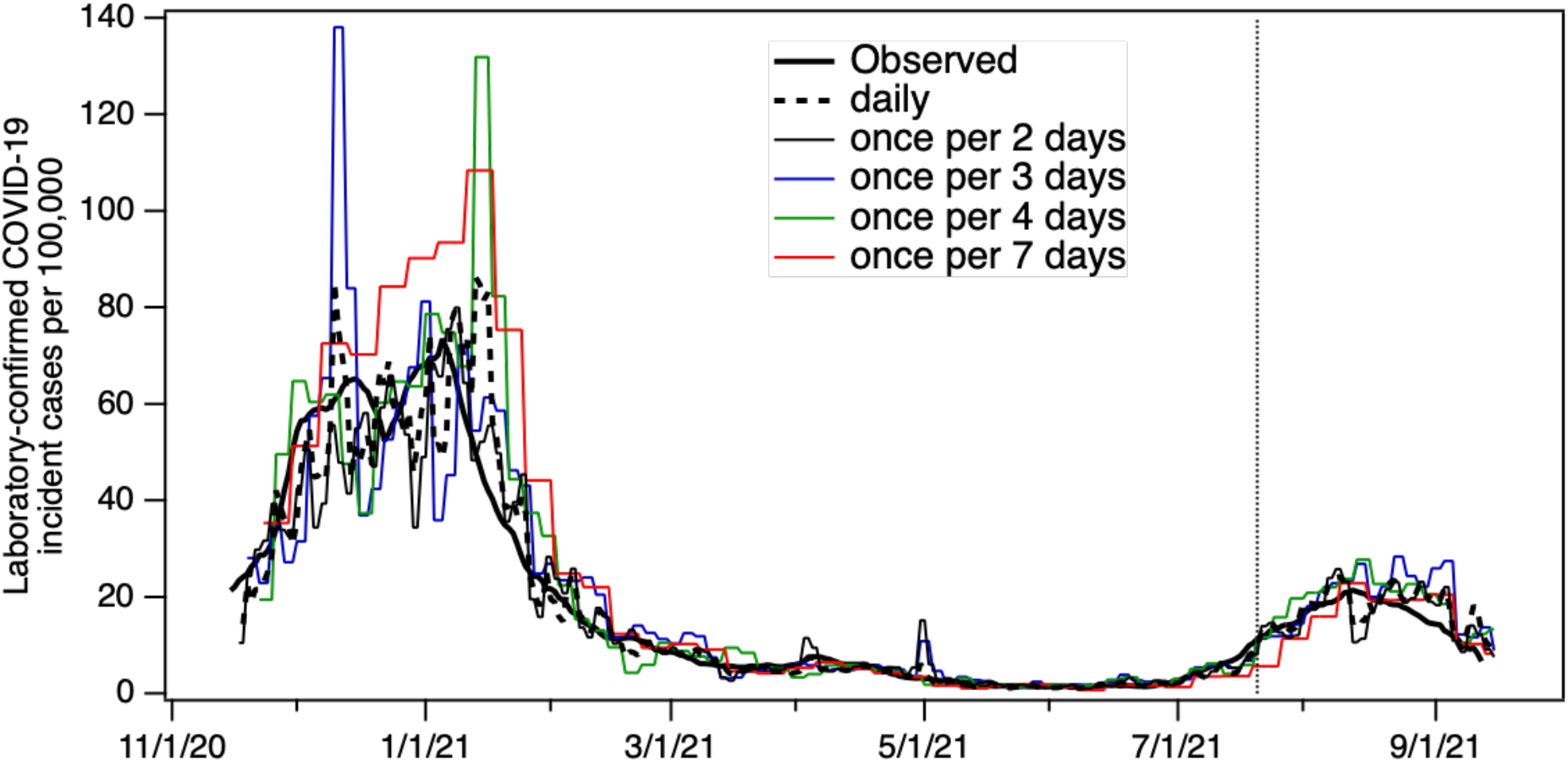
Predicted IR in-sample and out-of-sample prediction (after vertical line) using either daily N/PMMoV samples or with samples collected every other, third, and fourth day or weekly (trace for median in-sample RMSE presented). Results are shown for SJ; the other three POTWs are in Figure S1..

**Figure 4.**
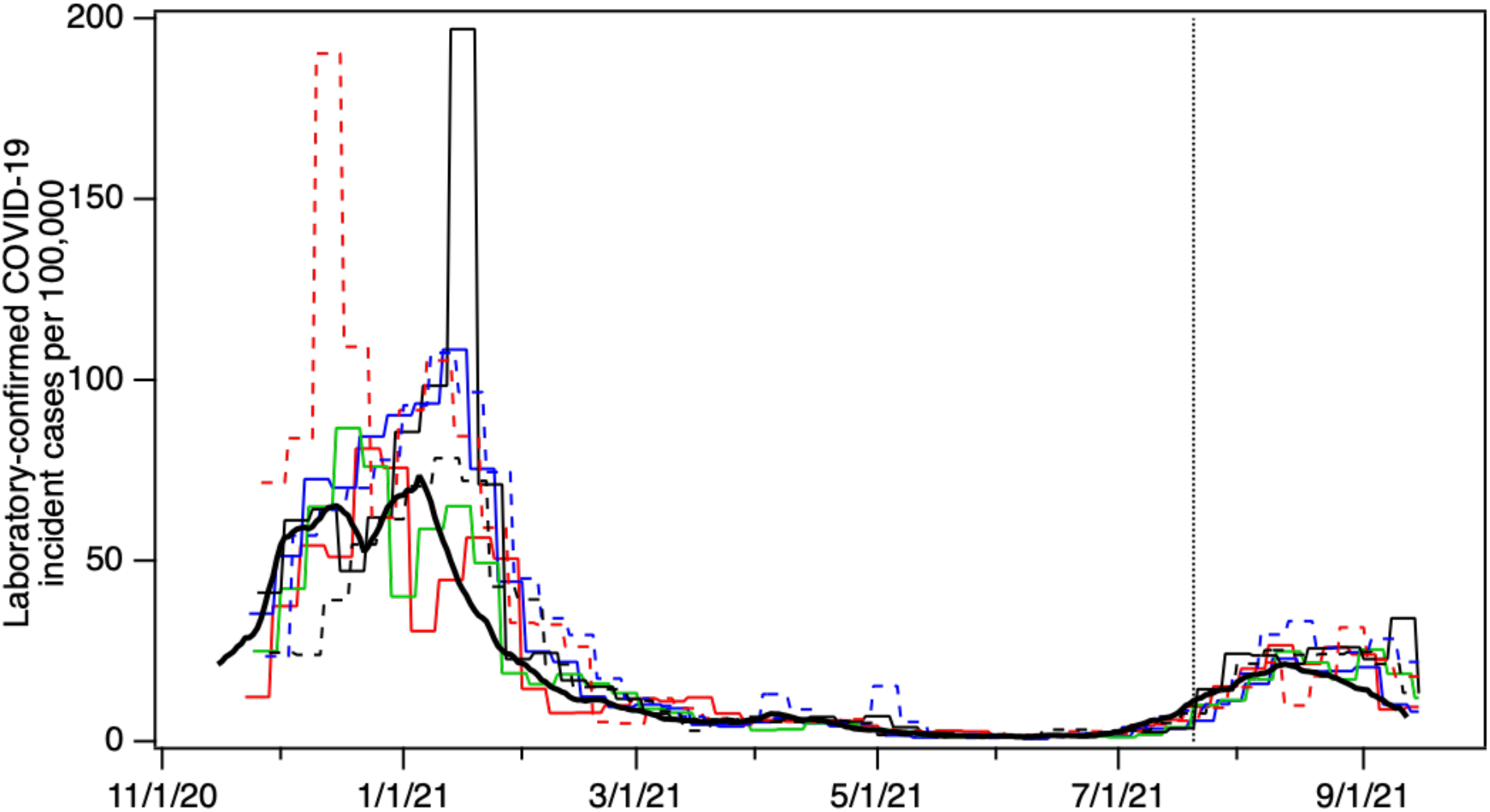
Predicted IR in-sample and out-of-sample prediction (after vertical line) for weekly sampling schemes for SJ. The different lines represent models generated with sampling conducted on different days of the week.

**Figure 5.**
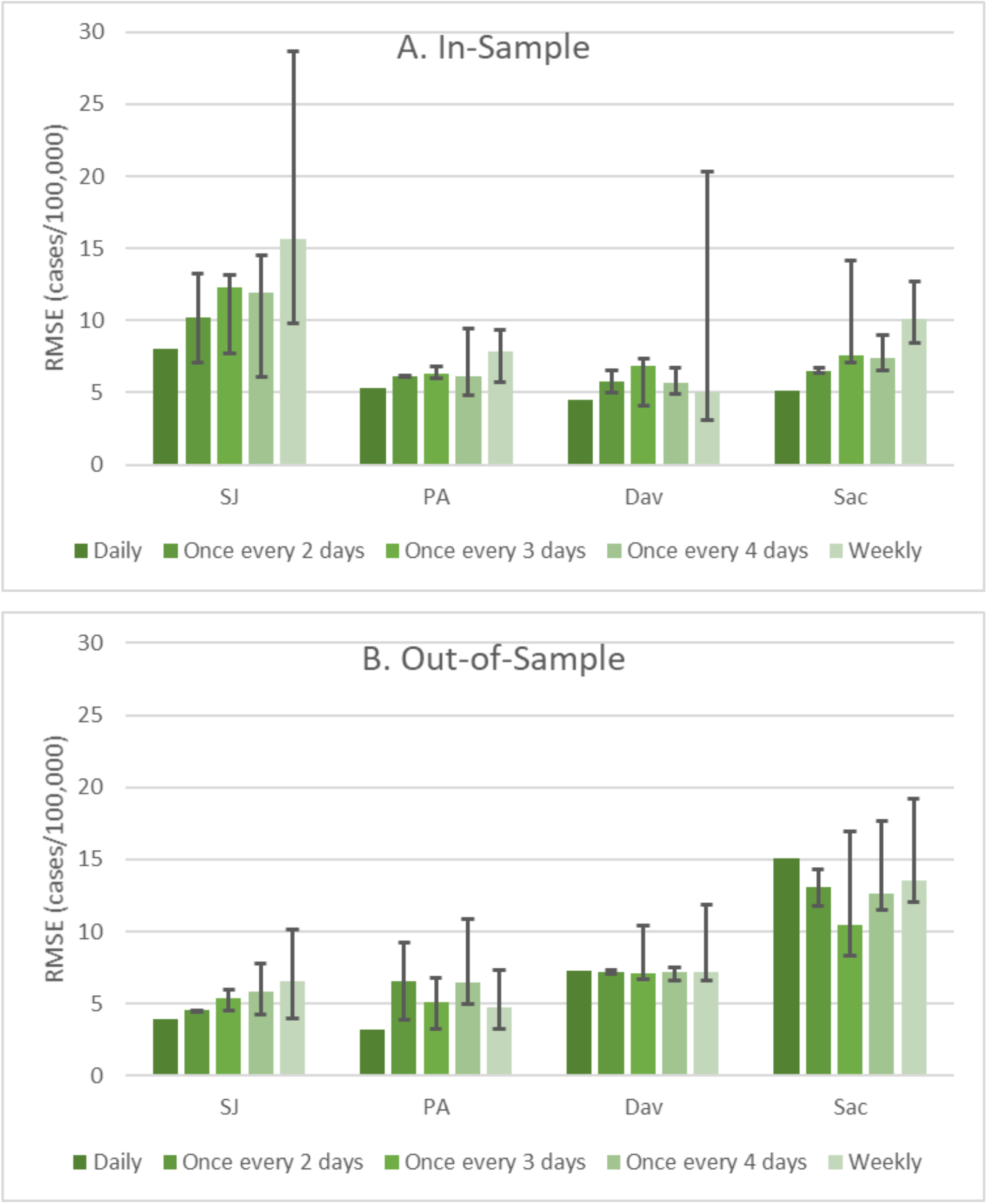
Comparison of in-sample and out-of-sample RMSE between DLMs fit using daily data and fit using reduced sampling frequencies (i.e., once every 2 days, once every 3 days … etc.). The predicted IRs on days missing N/PMMoV samples under reduced sampling frequency were estimated by freezing the most recent prediction. For reduced sampling frequency, median results shown with error bars capturing the range.

### 3.5 Comparison of sampling frequency

The RMSEs of the daily sampling model were less than the maximum RMSEs of the reduced frequency models across POTWs both in-sample and out-of-sample. Weekly sampling had the largest maximum RMSE of the models across POTWs both in-sample and out-of-sample (with the exceptions of in-sample Sac and out-of-sample PA). Weekly sampling had the greatest variability in RMSE with the widest range of roughly 9 to 28 cases/100,000 for SJ and followed by Dav with a range of 3 to 20 cases/100,000 during the in-sample period.

Generally, the largest RMSEs were estimated for the in-sample time period. When daily sampling was reduced to weekly sampling during the in-sample time period, the maximum RMSE increased by 20 cases/100,000 for SJ (followed by Dav, Sac, and PA with increases of 16, 8, and 4 cases/100,000, respectively). Whereas, for sampling frequencies of once every 2, 3, and 4 days, the maximum increase in RMSE from daily sampling was roughly 7 cases/100,000 for the in-sample period as well as the out-of-sample period. When sample frequency decreased to weekly sampling from the other reduced sampling frequencies, the maximum out-of-sample RMSE increased by < 2 cases/100,000 while the maximum in-sample RMSE increased up to 14 cases/100,000 (for SJ) when sampling was reduced to weekly.

## 4.0 Discussion

Sewage surveillance for SARS-CoV-2 RNA can provide daily, real-time information on disease burden in sewersheds.^1–3^ In this work, we investigate how the frequency of wastewater settled solids data affects daily predictions of laboratory-confirmed COVID-19 incidence rates generated using distributed lag models (DLMs) and show that while daily data produces the best estimates of incidence rate, reduced sampling frequencies that are likely to be more sustainable for long term programs can produce similar predictions.

To do this, we compared the predicted error, as estimated using the root mean square errors (RMSE), between models fit with daily wastewater data and models fit with wastewater data collected at lower frequencies (as low as once per week). The in- and out-of-sample RMSE was smallest for the model fit using the daily data across all 4 POTWs. Models fit using weekly wastewater data generally had the largest RMSE. The RMSEs of models fit using wastewater data collected at higher frequencies remained relatively small compared to those fit with weekly data.

This corroborates previous work that showed reduced sampling (2 x per week) maintained significant correlations between N1 concentration in influent and cases;^7^ resulted in significant associations using linear regression;^8^ and estimated similar effective reproductive number as daily sampling when sampled 3 x per week.^22^ Overall, the results of this study illustrate that real-time, daily prediction of incidence was possible using DLMs when sampling frequency was reduced.

### 4.2 Using a distributed lag model to estimate incidence rates

DLMs have been applied in epidemiology to relate exposure to a health outcome.^23–25^ However, the question remains if DLMs are useful for modeling COVID-19 incidence rates using wastewater data as explanatory variables. The predictive power of DLMs remains useful only if relationships hold over time, an assumption that may not hold for an infectious agent that acquires mutations and in a population that acquires immunity. Our results show strong out-of-sample prediction across multiple sites during the rise and subsequent dominance of the Delta variant using a model fit to an in-sample period when a variety of variants were present. We found that the variant did not have a significant influence on the predicted IR as % Delta was not a significant predictor across sites. However, additional work will need to be done to see if the relationship between SARS-CoV-2 RNA and incidence rates is robust to the emergence of other variants with distinct transmissibility or infection trajectories, like Omicron.

The coefficient estimates in the DLM indicate the effect of a temporary change in log_10_ N/PMMoV at time t on log_10_ IR in period t + lag. When log_10_ N/PMMoV was included in the model over consecutive days (e.g., lags = 0, 1, 2, 3, 3, 5, and 6), the effects on IR from changes in log_10_ N/PMMoV were similar across days, rather than pointing toward a specific lag as being important. However, when log_10_ N/PMMoV was included on non-consecutive days according to the reduced sampling frequency, the effects on IR from changes in log_10_ N/PMMoV were varied by day and site in a way that we were unable to explain.

### 4.3 DML limitations

This work illustrates limitations with DLM estimated via ordinary least squares. The residual plots illustrate that the selected DLM approach did not characterize all the structure of data. For example, the daily DLMs generally underpredicted during periods of peak incidence. Further, the residuals were highly autocorrelated. Modifications to capture the error structure (e.g., using weighted least squares regression) could further improve model performance. Multicollinearity was also present in the regressors, particularly for the data collected at higher sampling frequency, which can lead to unreliable coefficient estimates with large standard errors.^26^ However, this was not observed for the model fit to daily sampling data (Table 1 and Tables S2-S5). Multicollinearity can be addressed by putting constraints on the lagged effects such as adopting a spline function for the lag weights.^26^ Despite the flaws in the DLM approach selected, the out-of-sample prediction of IR was useful. Improvements to the models could further improve performance. In practice, the DLM can be continually updated over time by fitting to the entire dataset or a subset of interest. The use of DLM models relies on a fixed relationship between COVID-19 infections and wastewater concentrations of SARS-COV-2 RNA. During emergence of new variants, the relationship could change if shedding dynamics and loads vary. The models also rely on the existence of reliable COVID-19 infection data. Laboratory confirmed COVID-19 cases is a proxy for COVID-19 infections and may underpredict true infections particularly during periods of high positivity rates.^27^

### 4.4 Guidance on sampling frequency

When the primary use case for wastewater surveillance is to develop a predictive model that provides robust estimates of incidence rate over different phases of the pandemic, evidence from this study supports reduced sampling down to once every 4 days. However, this analysis does not consider the timeliness of these estimates or their use for trend analysis or variant identification and emergence. In addition, during periods of rapid changes in incidence or particularly high levels of disease, public health stakeholders may need daily information about outbreak dynamics to inform response. Frequent wastewater testing updates may be especially important due to reduced testing capacity or significant use of rapid antigen testing results that are not reported to public health during a surge. Aside from estimating incidence and identifying outbreak trends, an important use case of wastewater monitoring is also to identify the introduction of a variant into a population. During periods of suspected variant introduction, daily wastewater testing for variants may be desirable to identify this introduction in a timely manner, even if less frequent monitoring would serve the need to estimate overall incident rates at the time.

## 5.0 Conclusion

Distributed lag models had strong out-of-sample predictive power for COVID-19 incidence rates using wastewater solids surveillance of SARS-CoV-2 N genes (with lags of 0 to 3 days) for California sewersheds. The DLMs captured the rise and fall of cases during the introduction of the Delta variant using a model fit to data during a time period with a mixture of variants represented. When sampling frequency was reduced and out-of-sample prediction was made using a sampling frequency of once every 4 days or more, the DLMs provided real-time predicted incidence with low error. When reduced to weekly sampling frequency, the DLMs had variable predictive performance. Research shows that reduced frequency sampling programs which may be sustainable for long term monitoring can effectively be used to provide estimates of COVID-19 incidence; if capacity exists for increased testing frequency, there may be times in which more frequent testing would be desirable to either provide more frequent updates to public health or detect the early introduction of a new variant.

## Data Availability

Data are available as indicated in the paper through the Stanford Digital Repository.

## Acknowledgements

This work was funded by a gift from the CDC Foundation. We thank the California Department of Public Health COVID-19 Wastewater Surveillance, Epidemiology and Data teams for their help with COVID-19 incidence data. We thank Michael Balliet, Dr. Pamela Stoddard and Dr. George Han at County of Santa Clara Public Health Department for their continued support of the project, provision of case data, and public health leadership. Numerous people contributed to sample collection including Srividhya Ramamoorthy (Sac), Michael Cook (Sac), Ursula Bigler (Sac), James Noss (Sac), Lisa C. Thompson (Sac), Payak Sarkar (SJ), Noel Enoki (SJ), and Amy Wong (SJ), Karin North (PA), Armando Guizar (PA), and Jeromy Miller (Dav).

## Supporting Information

Tables S1-S7 show model outputs for all POTWs, Figure S1 shows data and predictions for all POTW, Figures S2-S5 show model residuals.

