## Supporting Information for "SARS-CoV-2 RNA wastewater settled solids surveillance frequency and impact on predicted COVID-19 incidence using a distributed lag model"

for

Table S1. SJ model fit comparison for daily sampling for dependent variable $log_{10}$IR (i.e., $log_{10}$ 7-day smoothed cases/population)

| Predictor | Coefficient Estimate (St. Error) | | |
|---|---|---|---|
| | Eq 1 | Eq 2 U=3 | Eq 2 U=6 |
| $log_{10}$ N/PMMoV $_0$ | 0.84*** | 0.25*** | 0.19*** |
| | (0.02) | (0.03) | (0.02) |
| $log_{10}$ N/PMMoV $_1$ | | 0.22*** | 0.16*** |
| | | (0.03) | (0.03) |
| $log_{10}$ N/PMMoV $_2$ | | 0.23*** | 0.15*** |
| | | (0.03) | (0.03) |
| $log_{10}$ N/PMMoV $_3$ | | 0.22*** | 0.13*** |
| | | (0.03) | (0.02) |
| $log_{10}$ N/PMMoV $_4$ | | | 0.10*** |
| | | | (0.03) |
| $log_{10}$ N/PMMoV $_5$ | | | 0.09*** |
| | | | (0.03) |
| $log_{10}$ N/PMMoV $_6$ | | | 0.07*** |
| | | | (0.03) |
| % Delta $_t$ | | | |
| Constant | -0.29*** | 0.03** | -0.07* |
| | (0.09) | (0.05) | (0.04) |
| BIC | -1.56 | -4.3 | -4.58 |
| Adjusted $R^2$ | 0.89 | 0.96 | 0.97 |
| Residual Std. Error | 0.20 | 0.11 | 0.10 |

∗ p<0.1; ∗∗ p<0.05; ∗∗∗ p<0.01

Table S2. PA model fit comparison for daily sampling for dependent variable $log_{10}$IR (i.e., $log_{10}$ 7-day smoothed cases/population)

| | Coefficient Estimate (St. Error) | | |
|---|---|---|---|
| Predictor | Eq 1 | Eq 2 U=3 | Eq2 U=6 |
| $log_{10}$ N/PMMoV $_0$ | 0.62*** | 0.18*** | 0.14*** |
| | (0.02) | (0.03) | (0.02) |
| $log_{10}$ N/PMMoV $_1$ | | 0.18*** | 0.14*** |
| | | (0.03) | (0.02) |
| $log_{10}$ N/PMMoV $_2$ | | 0.17*** | 0.13*** |
| | | (0.03) | (0.02) |
| $log_{10}$ N/PMMoV $_3$ | | 0.15*** | 0.11*** |
| | | (0.03) | (0.02) |
| $log_{10}$ N/PMMoV $_4$ | | | 0.07*** |
| | | | (0.02) |
| $log_{10}$ N/PMMoV $_5$ | | | 0.06*** |
| | | | (0.02) |
| $log_{10}$ N/PMMoV $_6$ | | | 0.06** |
| | | | (0.02) |
| % Delta $_t$ | | | |
| Constant | -1.31*** | -0.99*** | -0.90*** |
| | (0.02) | (0.07) | (0.05) |
| BIC | -1.34 | -3.52 | -3.87 |
| Adjusted $R^2$ | 0.80 | 0.91 | 0.92 |
| Residual Std. Error | 0.24 | 0.16 | 0.14 |

∗ p<0.1; ∗∗ p<0.05; ∗∗∗ p<0.01

Table S3. Dav model fit comparison for daily sampling for dependent variable $\log_{10}$IR (i.e., $\log_{10}$ 7-day smoothed cases/population)

| Predictor | Coefficient Estimate (St. Error) | | |
|---|---|---|---|
| | Eq 1 | Eq 2 U=3 | Eq 2 U=6 |
| $\log_{10}$ N/PMMoV $_0$ | $0.51^{***}$ | $0.18^{***}$ | $0.13^{***}$ |
| | (0.02) | (0.03) | (0.03) |
| $\log_{10}$ N/PMMoV $_1$ | | $0.12^{***}$ | $0.08^{***}$ |
| | | (0.03) | (0.03) |
| $\log_{10}$ N/PMMoV $_2$ | | $0.11^{***}$ | $0.06^{**}$ |
| | | (0.03) | (0.03) |
| $\log_{10}$ N/PMMoV $_3$ | | $0.17^{***}$ | $0.07^{**}$ |
| | | (0.03) | (0.03) |
| $\log_{10}$ N/PMMoV $_4$ | | | $0.08^{***}$ |
| | | | (0.03) |
| $\log_{10}$ N/PMMoV $_5$ | | | $0.08^{***}$ |
| | | | (0.03) |
| $\log_{10}$ N/PMMoV $_6$ | | | $0.11^{***}$ |
| | | | (0.03) |
| % Delta $_t$ | | | |
| Constant | $-1.97^{***}$ | $-1.66^{***}$ | $-1.49^{***}$ |
| | (0.09) | (0.08) | (0.06) |
| BIC | -0.6 | -3.05 | -3.26 |
| Adjusted $R^2$ | 0.75 | 0.85 | 0.88 |
| Residual Std. Error | 0.27 | 0.21 | 0.18 |

$*$ p<0.1; $**$ p<0.05; $***$ p<0.01

Table S4. Sac model fit comparison for daily sampling for dependent variable $\log_{10}$IR (i.e., $\log_{10}$ 7-day smoothed cases/population)

| Predictor | Coefficient Estimate (St. Error) Eq 1 | Eq 2 U=3 | Eq 2 U=6 |
|---|---|---|---|
| $\log_{10}$ N/PMMoV $_0$ | $0.61^{***}$ | $0.19^{***}$ | $0.15^{***}$ |
| | (0.02) | (0.03) | (0.03) |
| $\log_{10}$ N/PMMoV $_1$ | | $0.17^{***}$ | $0.14^{***}$ |
| | | (0.03) | (0.03) |
| $\log_{10}$ N/PMMoV $_2$ | | $0.14^{***}$ | $0.12^{***}$ |
| | | (0.03) | (0.03) |
| $\log_{10}$ N/PMMoV $_3$ | | $0.16^{***}$ | $0.11^{***}$ |
| | | (0.03) | (0.03) |
| $\log_{10}$ N/PMMoV $_4$ | | | $0.08^{***}$ |
| | | | (0.03) |
| $\log_{10}$ N/PMMoV $_5$ | | | $0.08^{**}$ |
| | | | (0.03) |
| $\log_{10}$ N/PMMoV $_6$ | | | 0.04 |
| | | | (0.03) |
| % Delta $_t$ | | | |
| Constant | $-1.38^{***}$ | $-1.20^{***}$ | $-0.92^{***}$ |
| | (0.07) | (0.05) | (0.06) |
| BIC | -1.87 | -4.63 | -4.21 |
| Adjusted $R^2$ | 0.84 | 0.93 | 0.90 |
| Residual Std. Error | 0.14 | 0.09 | 0.11 |

∗ p<0.1; ∗∗ p<0.05; ∗∗∗ p<0.01

Table S5. PA model fit comparison across sampling frequency for dependent variable $\log_{10}$IR (i.e., $\log_{10}$7-day smoothed cases/population)

| Predictor | Coefficient Estimate for predictors (St. Error) | | | | |
|---|---|---|---|---|---|
| | Daily | Once every 2 days | Once every 3 days | Once every 4 days | Weekly |
| $\log_{10}$ N/PMMoV $_{t-0}$ | 0.18*** (0.03) | 0.34*** (0.04) | 0.34*** (0.05) | 0.35*** (0.05) | 0.30*** (0.07) |
| $\log_{10}$ N/PMMoV $_{t-1}$ | 0.18*** (0.03) | | | | |
| $\log_{10}$ N/PMMoV $_{t-2}$ | 0.17*** (0.03) | 0.35*** (0.04) | | | |
| $\log_{10}$ N/PMMoV $_{t-3}$ | 0.15*** (0.03) | | 0.35*** (0.05) | | |
| $\log_{10}$ N/PMMoV $_{t-4}$ | | | | 0.36*** (0.05) | |
| $\log_{10}$ N/PMMoV $_{t-7}$ | | | | | 0.37*** (0.07) |
| $\alpha$ | -0.99*** (0.07) | -0.94*** (0.10) | -0.95*** (0.14) | -0.84*** (0.12) | -1.13*** (0.29) |
| Adjusted $R^2$ | 0.91 | 0.90 | 0.88 | 0.93 | 0.77 |

∗ p<0.1; ∗∗ p<0.05; ∗∗∗ p<0.01

Table S6. Dav model fit comparison across sampling frequency for dependent variable $log_{10}$IR (i.e., $log_{10}$7-day smoothed cases/population)

| Predictor | Coefficient Estimate for predictors (St. Error) | | | | |
|---|---|---|---|---|---|
| | Daily | Once every 2 days | Once every 3 days | Once every 4 days | Weely |
| $log_{10}$ N/PMMoV $_{0}$ | 0.18*** (0.03) | 0.25*** (0.04) | 0.31*** (0.05) | 0.20*** (0.06) | 0.22*** (0.07) |
| $log_{10}$ N/PMMoV $_{t-1}$ | 0.12*** (0.03) | | | | |
| $log_{10}$ N/PMMoV $_{t-2}$ | 0.11*** (0.03) | 0.28*** (0.04) | | | |
| $log_{10}$ N/PMMoV $_{t-3}$ | 0.17*** (0.03) | | 0.29*** (0.05) | | |
| $log_{10}$ N/PMMoV $_{t-4}$ | | | | 0.28*** (0.06) | |
| $log_{10}$ N/PMMoV $_{t-7}$ | | | | | 0.32*** (0.07) |
| $\alpha$ | -1.66*** (0.08) | -1.88*** (0.12) | -1.51*** (0.15) | -2.07*** (0.18) | -1.71*** (0.22) |
| Adjusted $R^2$ | 0.85 | 0.77 | 0.81 | 0.73 | 0.83 |

∗ p<0.1; ∗∗ p<0.05; ∗∗∗ p<0.01

Table S7. Sac model fit comparison across sampling frequency for dependent variable $log_{10}$IR (i.e., $log_{10}$ 7-day smoothed cases/population)

| Predictor | Coefficient Estimate for predictors (St. Error) | | | | |
|---|---|---|---|---|---|
| | Daily | Once every 2 days | Once every 3 days | Once every 4 days | Weely |
| $log_{10}$ N/PMMoV $_0$ | 0.19*** | 0.25*** | 0.26*** | 0.20** | 0.17 |
| | (0.03) | (0.07) | (0.08) | (0.10) | (0.12) |
| $log_{10}$ N/PMMoV $_{t-1}$ | 0.17*** | | | | |
| | (0.03) | | | | |
| $log_{10}$ N/PMMoV $_{t-2}$ | 0.14*** | 0.36*** | | | |
| | (0.03) | (0.07) | | | |
| $log_{10}$ N/PMMoV $_{t-3}$ | 0.16*** | | 0.45*** | | |
| | (0.03) | | (0.08) | | |
| $log_{10}$ N/PMMoV $_{t-4}$ | | | | 0.44*** | |
| | | | | (0.10) | |
| $log_{10}$ N/PMMoV $_{t-7}$ | | | | | 0.48*** |
| | | | | | (0.11) |
| $\alpha$ | -1.20*** | -1.30*** | -0.91*** | -1.21*** | -1.14*** |
| | (0.05) | (0.14) | (0.19) | (0.17) | (0.26) |
| Adjusted $R^2$ | 0.93 | 0.76 | 0.77 | 0.82 | 0.79 |

∗ $p<0.1$; ∗∗ $p<0.05$; ∗∗∗ $p<0.01$

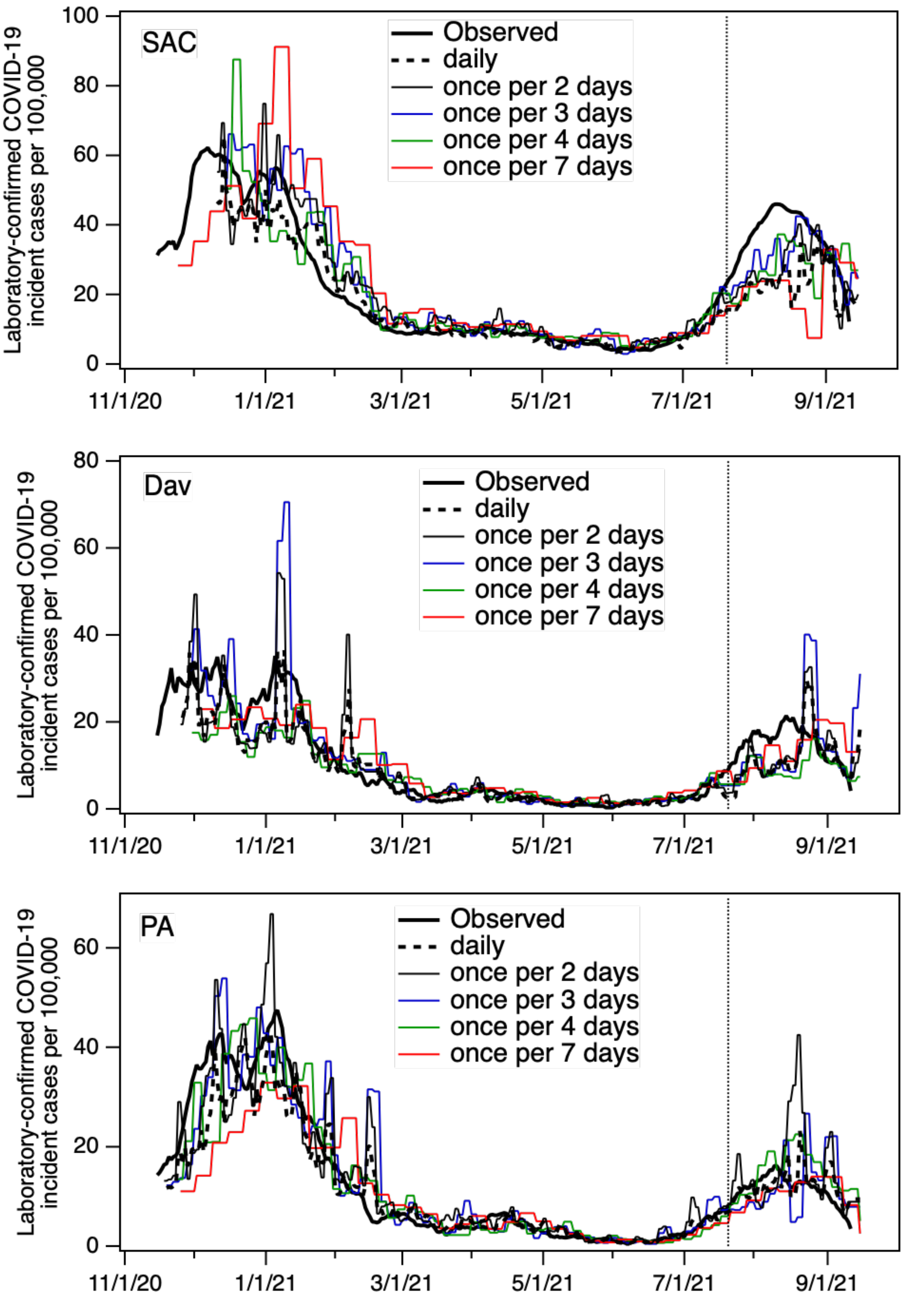


Figure S1. Predicted IR in-sample and out-of-sample prediction (after vertical line) using either daily N/PMMoV samples or with samples collected every other, third, and fourth day or weekly (trace for median in-sample RMSE presented). SJ presented in main text.

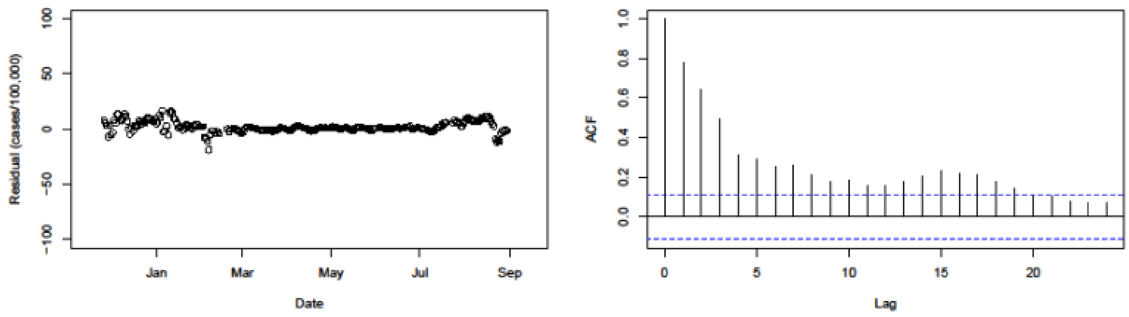


Figure S2. Residual plot and Autocorrelation of residual plot Eq 2 U = 3 for daily sampling Dav

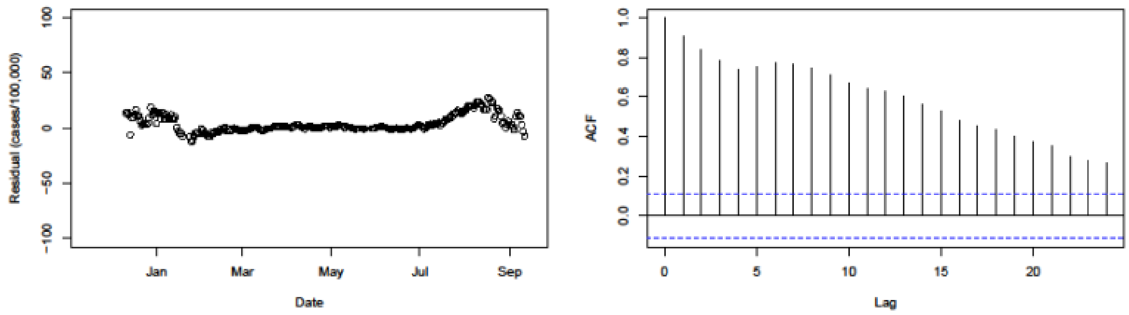


Figure S3. Residual plot and Autocorrelation of residual plot Eq 2 U = 3 for daily sampling Sac

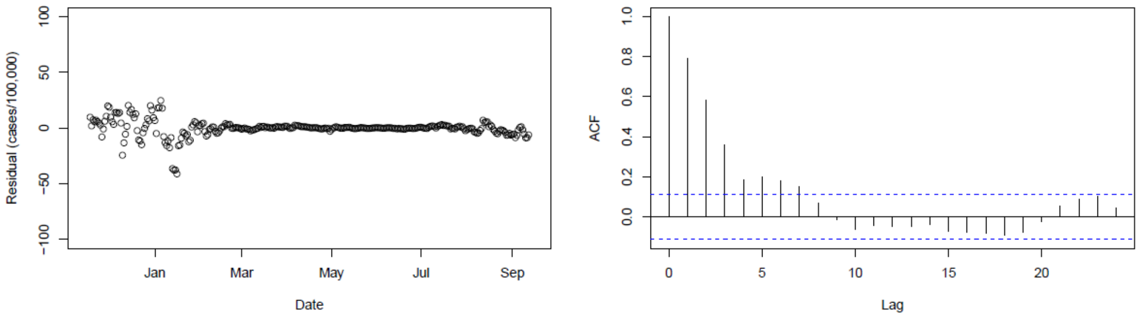


Figure S4. Residual plot and Autocorrelation of residual plot Eq 2 U = 3 for daily sampling SJ

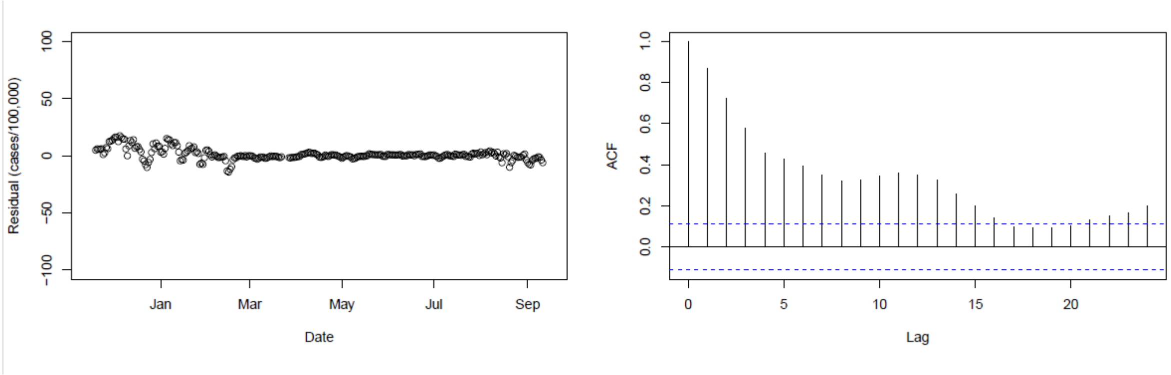


Figure S5. Residual plot and Autocorrelation of residual plot Eq 2 U = 3 for daily sampling PA
